# Sustained reductions in life-threatening invasive bacterial diseases during the first two years of the COVID-19 pandemic: analyses of prospective surveillance data from 30 countries participating in the IRIS Consortium

**DOI:** 10.1101/2022.12.16.22283251

**Authors:** David Shaw, Raquel Abad, Zahin Amin-Chowdhury, Adriana Bautista, Desiree Bennett, Karen Broughton, Bin Cao, Carlo Casanova, Eun Hwa Choi, Yiu-Wai Chu, Heike Claus, Juliana Coelho, Mary Corcoran, Simon Cottrell, Robert Cunney, Lize Cuypers, Tine Dalby, Heather Davies, Linda de Gouveia, Ala-Eddine Deghmane, Walter Demczuk, Stefanie Desmet, Mirian Domenech, Richard Drew, Mignon du Plessis, Carolina Duarte, Helga Erlendsdóttir, Norman Fry, Kurt Fuursted, Thomas Hale, Desiree Henares, Birgitta Henriques-Normark, Markus Hilty, Steen Hoffmann, Hilary Humphreys, Margaret Ip, Susanne Jacobsson, Christopher Johnson, Jillian Johnston, Keith A Jolley, Aníbal Kawabata, Jana Kozakova, Karl G Kristinsson, Pavla Krizova, Alicja Kuch, Shamez Ladhani, Thiên-Trí Lâm, León María Eugenia, Laura Lindholm, David Litt, Martin CJ Maiden, Irene Martin, Delphine Martiny, Wesley Mattheus, Noel D McCarthy, Martha McElligott, Mary Meehan, Susan Meiring, Paula Mölling, Eva Morfeldt, Julie Morgan, Robert Mulhall, Carmen Muñoz-Almagro, David Murdoch, Joy Murphy, Martin Musilek, Alexandre Mzabi, Ludmila Novakova, Shahin Oftadeh, Amaresh Perez-Arguello, Maria Pérez-Vázquez, Monique Perrin, Malorie Perry, Benoit Prevost, Maria Roberts, Assaf Rokney, Merav Ron, Olga Marina Sanabria, Kevin J Scott, Carmen Sheppard, Lotta Siira, Vitali Sintchenko, Anna Skoczyńska, Monica Sloan, Hans-Christian Slotved, Andrew J Smith, Anneke Steens, Muhamed-Kheir Taha, Maija Toropainen, Georgina Tzanakaki, Anni Vainio, Mark PG van der Linden, Nina M van Sorge, Emmanuelle Varon, Sandra Vohrnova, Anne von Gottberg, Jose Yuste, Rosemeire Zanella, Fei Zhou, Angela B Brueggemann

## Abstract

**Background:** The Invasive Respiratory Infection Surveillance (IRIS) Consortium was established to assess the impact of the COVID-19 pandemic on invasive diseases caused by *Streptococcus pneumoniae, Haemophilus influenzae, Neisseria meningitidis* and *Streptococcus agalactiae*. Here we analyse the incidence and distribution of disease during the first two years of the pandemic.

**Methods:** Laboratories in 30 countries/territories representing five continents submitted case data from 2018-2021 to private projects within databases in PubMLST. The impact of COVID-19 containment measures on the overall number of cases was analysed, and changes in disease distributions by patient age and serotype/group were examined. Interrupted time series analyses quantified the impact of pandemic response measures and their relaxation on disease rates, and autoregressive integrated moving average models estimated effect sizes and forecasted counterfactual trends by hemisphere.

**Findings:** Overall, 116,841 cases were analysed: 76,481 (2018-2019, pre-pandemic) plus 40,360 (2020-2021, pandemic). During the pandemic there was a significant reduction in the risk of disease caused by *S pneumoniae* (risk ratio: 0.47; 95% confidence interval: 0.40-0.55), *H influenzae* (0.51; 0.40-0.66) and *N meningitidis* (0.26; 0.21-0.31), whereas no significant changes were observed for the non-respiratory-transmitted pathogen *S agalactiae* (1.02; 0.75-1.40). No major changes in the distribution of cases were observed when stratified by patient age or serotype/group. An estimated 36,289 (17,145-55,434) cases of invasive bacterial disease were averted during the first two years of the pandemic among IRIS participating countries/territories.

**Interpretation:** COVID-19 containment measures were associated with a sustained decrease in the incidence of invasive disease caused by *S pneumoniae, H influenzae* and *N meningitidis* during the first two years of the pandemic, but cases began to increase in some countries as pandemic restrictions were lifted.

**Research in context:** *Evidence before this study:* Early in the COVID-19 pandemic the IRIS Consortium reported a significant reduction in invasive disease due to respiratory-transmitted bacterial pathogens, which was associated with the implementation of COVID-19 stringency measures and changes in human social behaviour. All 26 countries/territories participating in IRIS at the time experienced a significant reduction in infections between January and May 2020, compared with the previous two years. In particular, *S pneumoniae* infections decreased by 68% at four weeks after COVID-19 containment measures were imposed, and by 82% at eight weeks.

*Added value of this study:* These new data from the expanded IRIS Consortium across 30 countries/territories demonstrated a sustained reduction in invasive disease throughout the first two years of the COVID-19 pandemic. Using time series modelling, we estimated that over 36,000 cases of invasive bacterial disease were averted in 2020-2021 among the countries participating in IRIS; however, minor increases in disease in the latter half of 2021 require close monitoring to understand the nature of re-emerging cases.

*Implications of all the available evidence:* Future epidemics and pandemics will occur, and we need to understand not only the pathogen that is directly responsible for the pandemic, but also that population-level responses to an epidemic or pandemic more broadly affect overall human health and other microbes. IRIS provides evidence for the effects of such public health responses on severe invasive bacterial infections across many countries. Moreover, these IRIS data provide a better understanding of microbial transmission, will inform vaccine development and implementation, and can contribute to healthcare service planning and provision of policies.

## Introduction

Three of the most common causes of invasive bacterial disease are *Streptococcus pneumoniae, Haemophilus influenzae* and *Neisseria meningitidis*, and young children, adolescents and older adults are at greatest risk of disease. All three bacterial species colonise the oro- or nasopharynx of healthy individuals and all are transmitted person-to-person via respiratory droplets.

The Global Burden of Diseases, Injuries, and Risk Factors Study (GBD) estimated that, in 2019, *S pneumoniae* was the leading bacterial cause of death among children less than five years of age worldwide (225,000 deaths, 95% uncertainty interval (UI) 180,000–281,000). It was also the leading cause of lower respiratory infections (653,000 deaths, 95% UI, 553,000-777,000) and meningitis (44,500 deaths, 95% UI, 34,700-59,800) among people of all ages, and led to 40.3 million (95% UI, 32.8-50.0 million) years of life lost globally (1). The same report estimated a total of 101,000 deaths (95% UI, 82,800-124,000) due to *H influenzae*, the majority of which were due to lower respiratory infections, and 141,000 estimated deaths (95% UI, 96,800-203,000) from bloodstream infections and meningitis caused by *N meningitidis* (1). *N meningitidis* is a global pathogen but a particular public health problem in Africa since it is a cause of meningitis epidemics, both within and outside the ‘meningitis belt’ (2). *S pneumoniae* and *H influenzae* are also among the most common causes of deaths associated with infections caused by bacteria that are resistant to antimicrobials (3).

The Invasive Respiratory Infection Surveillance (IRIS) Consortium, an international network of microbiology laboratories in 30 countries and territories, was established early in 2020 in response to the pandemic caused by SARS-CoV-2 and concerns about the potential for increased post-viral secondary bacterial infections (4). The main aim of the IRIS Consortium is to investigate the incidence of invasive diseases caused by *S pneumoniae, H influenzae* and *N meningitidis*. Invasive infections due to these bacterial species are legally notifiable to public health registries in the majority of countries participating in IRIS.

Previously, we reported significant reductions in the incidence of diseases caused by all three bacteria early in the pandemic and demonstrated that these reductions were associated with the timing and stringency of COVID-19 containment measures (4-9). A subset of laboratories also submitted data for infections caused by *Streptococcus agalactiae*, which is a major cause of neonatal invasive disease (1,10) but is not transmitted via the respiratory route. *S agalactiae* was included as a comparator organism to assess the stability of routine disease surveillance during the pandemic. There was no change in the incidence of invasive *S agalactiae* infections in the early months of the pandemic, suggesting that any disruptions to routine laboratory surveillance during the COVID-19 pandemic were minor and did not explain the observed reductions in diseases caused by *S pneumoniae, H influenzae* and *N meningitidis* (4).

Here, we report an expanded analysis to include disease data for all four bacterial species in the two years pre-COVID-19 (2018-2019) and the first two years of the pandemic (2020-2021). Four countries were added to the IRIS Consortium since our first publication, which expanded the geographical coverage of IRIS to 30 countries/territories across five continents. We collected data on patient age and bacterial serotype/group to assess epidemiological changes that may have implications for disease burden and vaccination programmes. We quantified the effect of COVID-19 restrictions on the four pathogens under investigation, utilised time series modelling techniques to analyse changes in disease during the first two years of the pandemic, and estimated the number of cases averted. Our study also assessed the incidence of disease by patient age and serotype/group, to investigate whether the epidemiology of invasive disease had changed during 2020-2021 as compared to the pre-pandemic years.

## Methods

### Study design and participants

National reference and expert microbiology laboratories in 30 countries submitted data on confirmed cases of invasive infections (within a normally sterile site) caused by one or more of the four bacteria under investigation. Australia, Colombia, Greece, and Paraguay joined IRIS since the initial establishment of the consortium. All IRIS participating laboratories provided national reference data apart from Australia (New South Wales only) and China (one Beijing hospital only). Data were collected for patients of all ages except in South Korea, where only data from patients less than 16 years of age were available for analyses.

### Data collection

No patient-identifiable data were submitted to IRIS. In the originating laboratories, bacteria from clinical samples were primarily recovered and identified by standard microbiological culture methods and occasionally by PCR testing. Invasive disease cases identified from 1 January 2018 to 2 January 2022 (the end of ISO [International Organization for Standardization] week 52 for 2021), were included in the current analyses. The PubMLST suite of databases were used to collect and manage IRIS data, and a private project only accessible to IRIS participants was used for each of the four organisms. At a minimum, the specimen sampling date, patient age and serotype/group were submitted for each case except where data protection rules in a country prevented the submission of patient age. Study data were entered by IRIS participants or the database curators (ABB, KAJ, and DS). Automated data integrity checks were applied before data upload, and all IRIS data were manually checked by the curators for data consistency; any discrepant or missing data were queried and resolved with the submitting laboratory.

Google COVID-19 Community Mobility Reports (CCMR) are anonymised, within-country mobile device location history data that capture the movement of people in six categories, including time spent in workplaces and residential areas. Google CCMR data are calculated as a daily percentage change from the baseline day, which was the median value between 3 January and 6 February 2020. In our first paper we used the Google CCMR data to estimate the week when each country first implemented COVID-19 containment measures. We used those estimates in the current analyses, and for the four additional countries the week of implementation was calculated as described previously (4).

The stringency of each country’s COVID-19 containment measures was quantified using the Oxford Blavatnik COVID-19 Government Response Tracker (OxCGRT; 11). This stringency index combines nine indicators that are tracked daily: school, workplace and public transport closures; public event cancellations; gathering restrictions; stay at home requirements; internal movement restrictions; international travel controls; and public information campaigns. A composite stringency index variable between 0-100 is calculated and is available for download on the OxCGRT website. For our analyses, the daily stringency index was converted into an ISO weekly index by taking the mean stringency index metric for that week. Cumulative weekly case counts for each organism were plotted against the weekly stringency index for each country and organism.

### Time series analysis and decomposition

Case counts were summed by month to generate country- and organism-specific time series for 2018-2021. Monthly case totals were used to improve the overall model fit and accommodate the 2020 leap year (ie 53 weeks). A second time series analysis was performed for the *S pneumoniae, H influenzae* and *N meningitidis* datasets to account for potential seasonal differences, whereby case counts were pooled by countries residing in the Northern or Southern hemispheres, respectively. *S agalactiae* data were only collected from countries in the Northern hemisphere.

### Interrupted time series analysis

Seasonal autoregressive integrated moving average (ARIMA) models were used to quantify the impact of COVID-19 containment measures on the incidence of invasive bacterial disease, and to generate counterfactual trends with prediction intervals for each of the four pathogens. Although it would have been desirable to fit separate ARIMA models by country, the dataset in most countries was too small for this to be implemented, and residual autocorrelation and negative case counts produced after model fitting precluded this approach.

ARIMA models took the simplified form of:

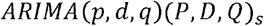

where *p* = non-seasonal autoregressive (AR) order, *d* = non-seasonal differencing, *q* = non-seasonal moving average (MA) order, *P* = seasonal AR order, *D* = seasonal differencing, *Q* = seasonal MA order, and *s* = time span of repeating seasonal pattern (number of observations in a year), 12 months.

Box-Jenkins methodology was applied when building the ARIMA model and both manual and automated methods were used to select the final models (12). Each time series was assessed for stationarity using the augmented Dickey-Fuller unit root test, which tested the null hypothesis of non-stationarity (12). None of the individual time series analysed in this study were non-stationary so no adjustments were required.

Manual model identification and coefficient estimation were performed, accounting for any seasonal pattern in the time series and utilising the autocorrelation function (ACF) and the partial autocorrelation function (PACF) to decide on the potential inclusion of and number of autoregressive and/or moving average components (order selection). Maximum likelihood estimation (MLE) methods were used to select coefficient values. Akaike information criterion (AIC) and Bayesian information criterion (BIC) values were used to select the best fitting model, with a preference for the most parsimonious model (12,13).

To measure the impact of COVID-19 containment measures, step and slope variables were included as regressors in the final ARIMA models: step (0 before containment measure implementation, 1 thereafter); and slope (0 before containment measure implementation, +1 for each month thereafter). Based on Google CCMR data, the step variable switched from 0 to 1 from March 2020 onwards for all countries. The final ARIMA models were used to produce a counterfactual prediction that assumed the COVID-19 pandemic did not occur, based on disease data from the two pre-pandemic years, which generated a mean monthly case estimate and 95% prediction interval (95% PI). The relative risk (RR) of invasive disease during the pandemic, and number of cases averted, were estimated.

### Meta-analysis

The RR estimates of disease and 95% confidence intervals (CI) in the Northern and Southern hemispheres were combined using an inverse-variance weighted, fixed effects meta-analysis along with random effects meta-analytic models to generate a pooled RR and 95% CI estimate. These models employed a restricted maximum likelihood approach for coefficient estimates. This approach was also applied to pool the results of the various sensitivity analyses.

### Sensitivity Analysis

For each of the four pathogens, segmented regression models were fitted to each country, stratified by age group and serotype/group. Negative binomial and quasi-Poisson generalised linear models were fitted to account for overdispersion of data, using population size as an offset and adjusting for seasonality, using month as a factor variable and Fourier terms. These models included a step change variable for implementation of containment measures as described above. We also included models that extrapolated a counterfactual trend from pre-pandemic data.

## Results

All 30 countries participating in IRIS submitted data on cases of *S pneumoniae* invasive disease, as represented by bacterial isolates and/or case reports submitted to each of the IRIS laboratories. Most countries also submitted *H influenzae* (n=24) and *N meningitidis* (n=21) invasive disease data, and nine countries submitted *S agalactiae* invasive disease data. Overall, the number of *S pneumoniae, H influenzae* and *N meningitidis* cases during 2020-2021 was approximately half the expected number each year as compared to pre-pandemic totals, but the number of *S agalactiae* cases was similar each year (Table 1).

**Table 1.** Overall number of invasive disease cases submitted to IRIS participating laboratories before (2018-2019) and during (2020-2021) the COVID-19 pandemic.

| Year | Total | *S pneumoniae* | *H influenzae* | *N meningitidis* | *S agalactiae* |
| --- | --- | --- | --- | --- | --- |
| 2018 | N | 30,533 | 3,510 | 2,302 | 1,702 |
|  | Median | 2,405 | 282 | 193 | 147 |
|  | IQR | 1,824-3,162 | 248-360 | 162-207 | 130-155 |
| 2019 | N | 30,606 | 3,697 | 2,228 | 1,883 |
|  | Median | 2,523 | 313 | 178 | 153 |
|  | IQR | 1,759-3,026 | 269-341 | 163-191 | 142-170 |
| 2020 | N | 15,501 | 2,120 | 976 | 1,886 |
|  | Median | 753 | 115 | 40 | 154 |
|  | IQR | 614-957 | 96-129 | 36-57 | 136-164 |
| 2021 | N | 15,306 | 2,117 | 550 | 1,904 |
|  | Median | 1,159 | 157 | 39 | 153 |
|  | IQR | 961-1,219 | 121-165 | 33-55 | 149-166 |

Note: median, monthly median value; N, total number of isolates per year; IQR, interquartile range; year, ISO year.

There was an inverse relationship between the stringency of the COVID-19 containment measures implemented in each country and the number of *S pneumoniae* cases reported to laboratories, ie as the stringency of containment measures decreased during 2021 in many countries, there were concomitant increases in *S pneumoniae* disease (Figure 1). Similar inverse relationships and increases in disease were observed for *H influenzae* and *N meningitidis* (Supplementary Figures 1 and 2), but not *S agalactiae* (Supplementary Figure 3).

**Figure 1.**
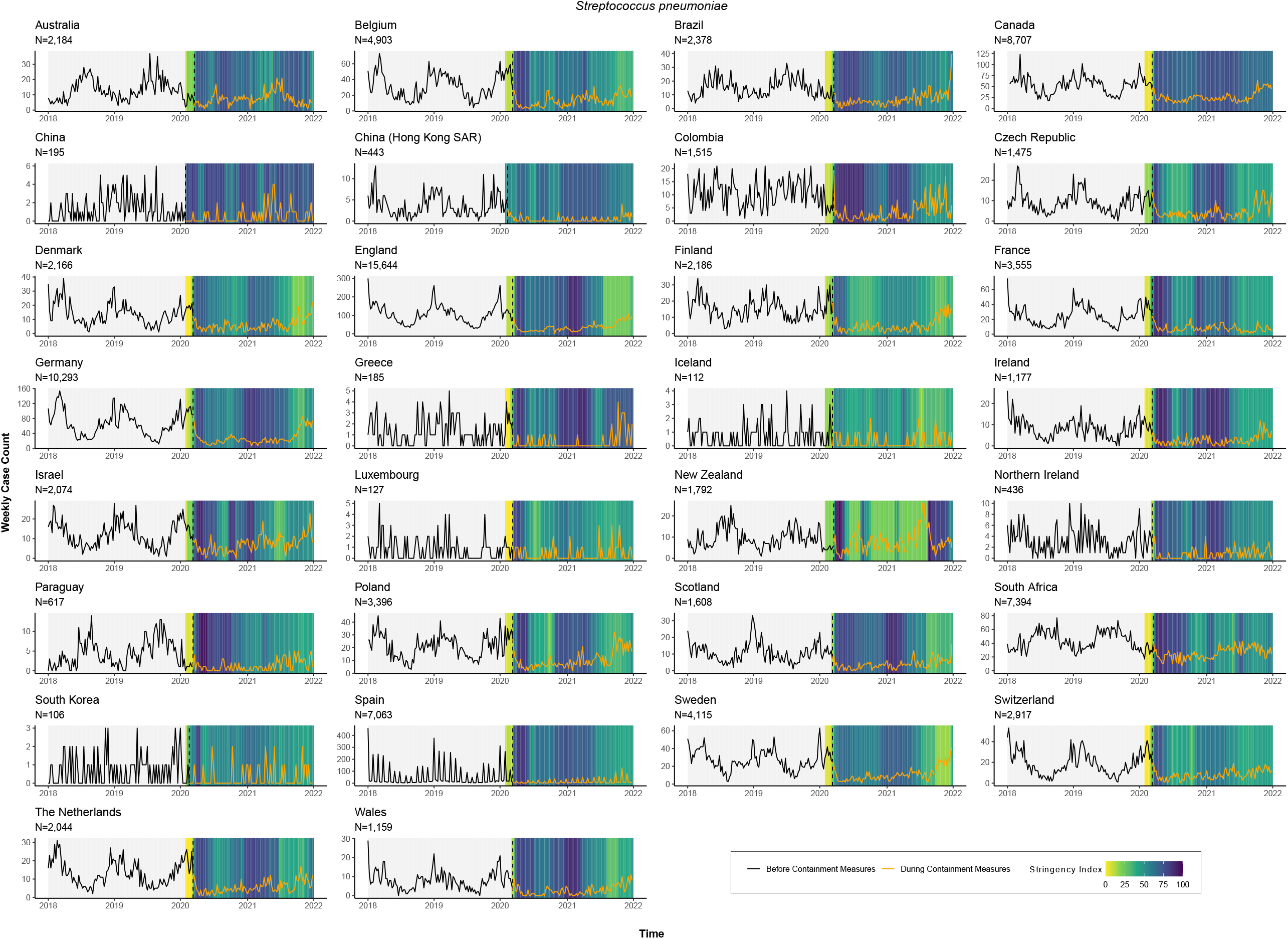
*S pneumoniae* invasive disease case counts. For each country, weekly invasive disease cases from 1 January 2018 to 2 January 2022 (four complete ISO years) were plotted against the weekly Oxford COVID-19 Government Response Tracker stringency index value in 2020-2021. The vertical hashed line indicates the week in which pandemic response measures were initiated in each country. Note that many of the Spanish sampling dates were submitted only by month and not day of sampling, so the sampling date was entered as the first day of the month if the actual sampling day was unavailable.

Time series analyses by Northern and Southern hemispheres for each of the bacterial species exhibited the overall reduction in diseases caused by *S pneumoniae, H influenzae* and *N* meningitidis, but not *S agalactiae*, during the pandemic (Figure 2). Notably, disease due to *S pneumoniae, H influenzae* and *N meningitidis* was increasing by the end of 2021. Google CCMR data were used to assess the point at which there was a step change that precipitated the reduction in disease within each country, which was from March 2020 (when the pandemic was declared) in both the Northern and Southern hemispheres (Figure 2).

**Figure 2.**
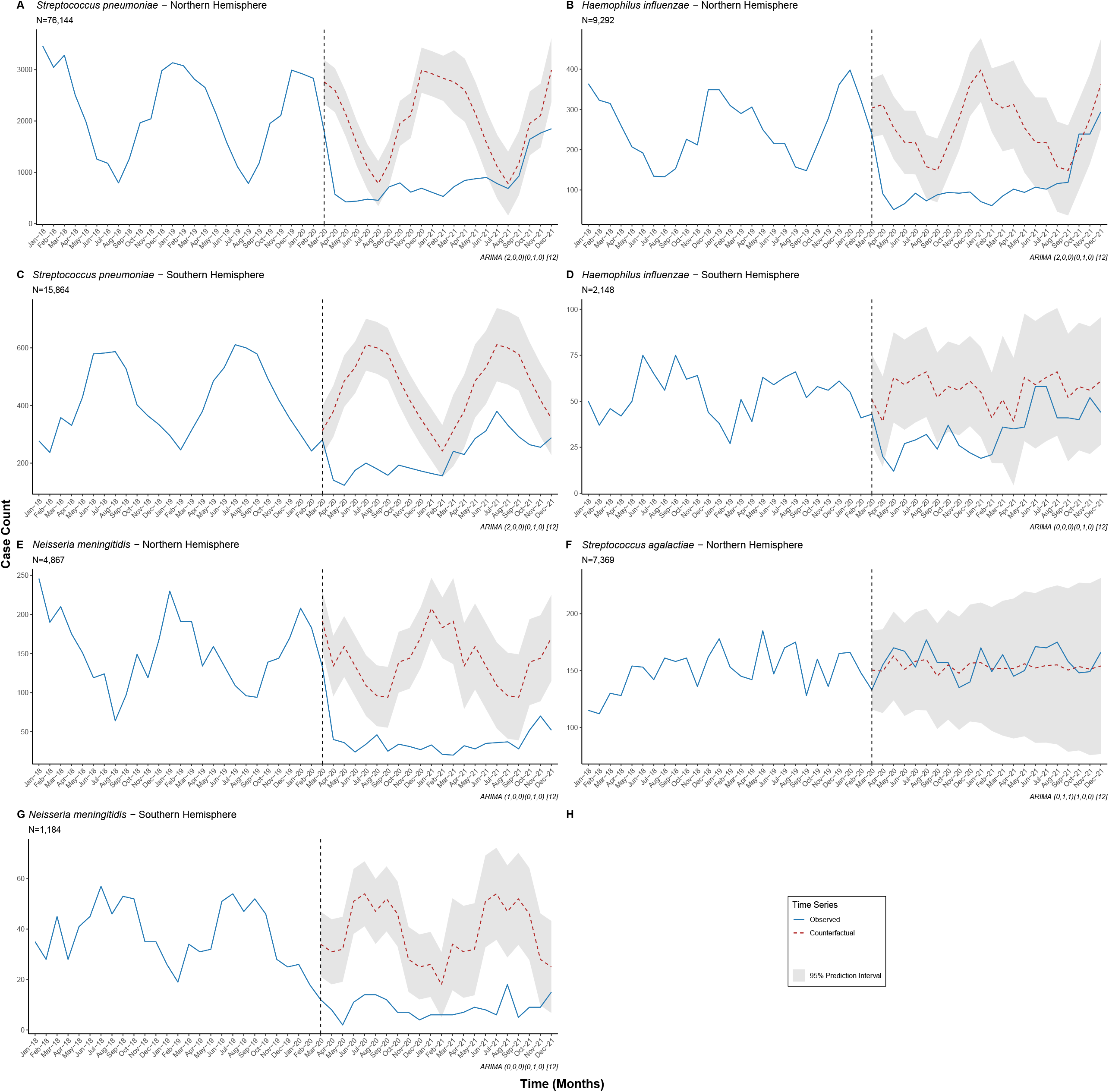
Interrupted time series analyses of invasive disease data by Northern and Southern hemispheres. Observed cases of invasive disease for each bacterial species (blue lines) were plotted against the counterfactual weekly number of cases predicted by the ARIMA models (red hashed lines) if the COVID-19 pandemic had not occurred. The vertical hashed line indicates the week in which pandemic response measures were initiated in each country. Note that *S agalactiae* data were only collected in the Northern hemisphere, and that data are plotted by weeks in the calendar year rather than ISO year.

The data were meta-analysed by hemisphere, the results of which demonstrated a significant reduction in the risk of invasive disease caused by *S pneumoniae* (RR 0.47, 95% CI, 0.40-0.55), *H influenzae* (RR 0.51, 95% CI, 0.40-0.66) and *N meningitidis* (RR 0.26, 95% CI, 0.21-0.31) but not *S agalactiae* (RR 1.02, 95% 0.75-1.40; Figure 3). Sensitivity analyses supported the use of the ARIMA model (Supplementary Figure 4), which estimated that 36,289 (17,145-55,434) cases were averted in these 30 countries during the first two years of the pandemic (Table 2).

**Figure 3.**
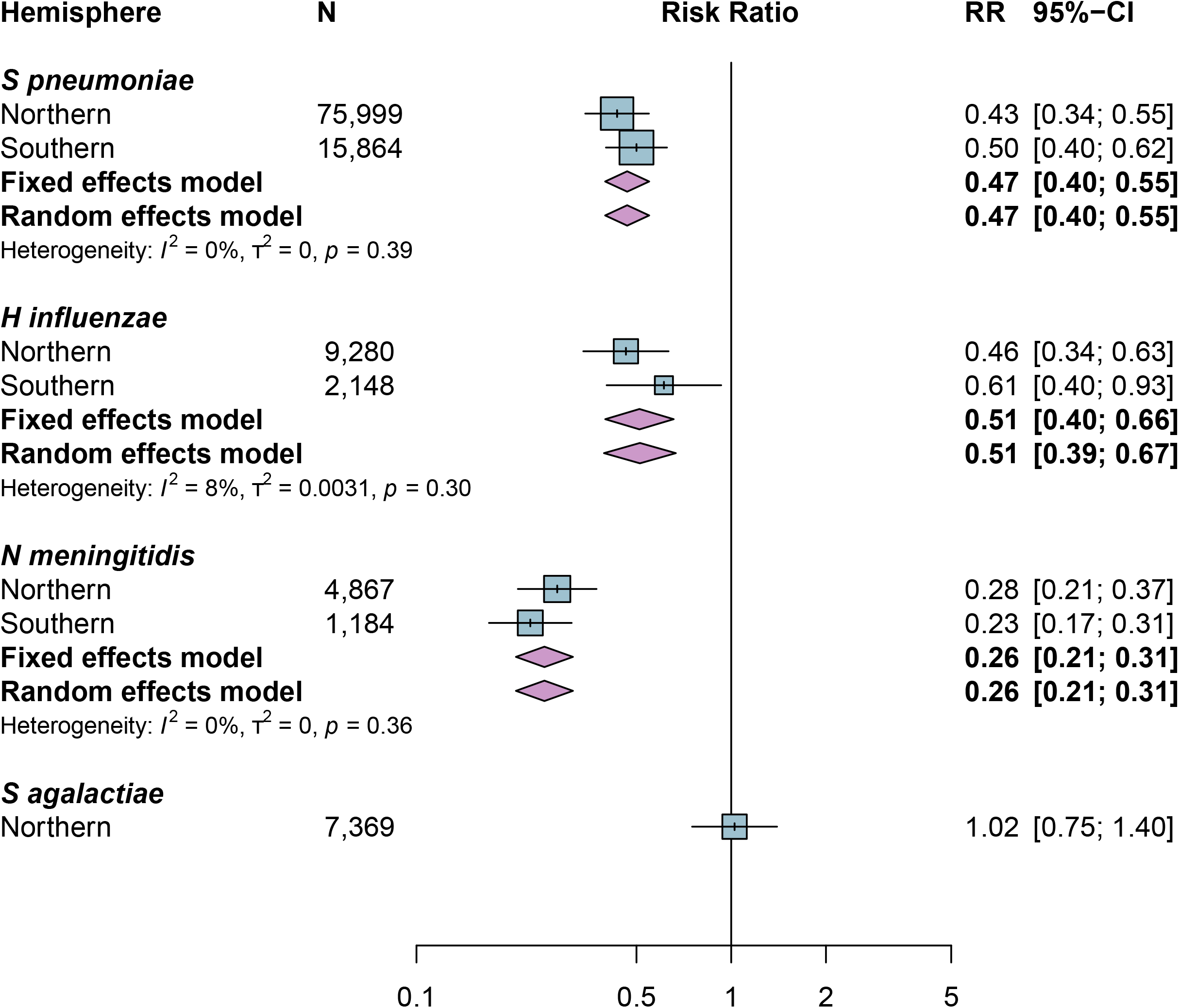
Risk of invasive disease during the pandemic for each bacterial species by hemisphere. Results of the meta-analysis are shown as fixed effects and random effects model estimates for each bacterial species. Note: RR, risk ratio; CI, confidence interval.

**Table 2.** Estimated number of invasive disease cases averted during the pandemic, 2020-2021, by hemisphere.

| Organism | Hemisphere | Cases averted | 95% CI |  |
| --- | --- | --- | --- | --- |
|  |  |  | lower | upper |
| *S pneumoniae* | Northern | 24,893 | 13,377 | 36,410 |
|  | Southern | 5,067 | 2,726 | 7,409 |
| *H influenzae* | Northern | 3,027 | 966 | 5,089 |
|  | Southern | 479 | -162 | 1,120 |
| *N meningitidis* | Northern | 2,258 | 1,240 | 3,275 |
|  | Southern | 649 | 312 | 986 |
| *S agalactiae* | Northern | -84 | -1,314 | 1,145 |

Note: CI, confidence interval

Data were then stratified by serotype/group and patient age. Among *S pneumoniae* cases, there were significant reductions among all major serotypes in 2020-2021, although cases of disease due to some serotypes were beginning to increase in the latter months of 2021 (Figure 2 and 4A). Case numbers in 2020-2021 were reduced in every age category and there were no major changes in the overall patterns of disease by age or serotype (Figure 4B).

**Figure 4.**
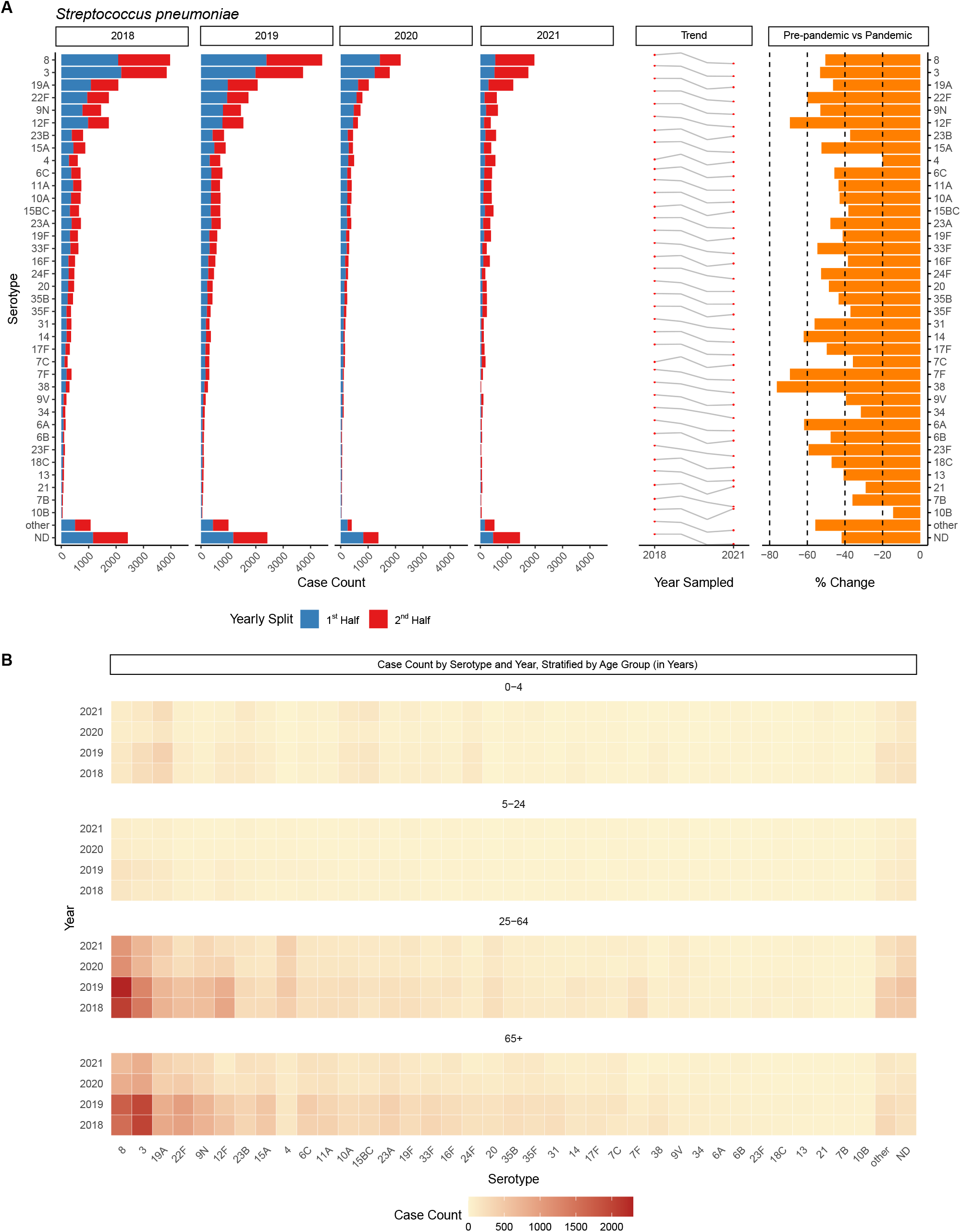
*S pneumoniae* invasive disease cases by serotype and patient age. Panel A: Distribution of serotypes responsible for 90% of all reported cases between 2018-2021, listed by case count, increasing or decreasing trend year-by-year, and percentage change of each serotype recovered in 2018-2019 compared to 2020-2021 (average number of cases each year, pre-pandemic vs pandemic). Panel B: Heat map depicting the number of cases of each serotype recovered per year and by age group. Note: serotype ND = not determined.

Stratification of *H influenzae* cases by serotype and patient age demonstrated a reduction in all serotypes except serotype b (Hib), which decreased in 2020 and then increased (P<0.01) at the end of 2021. However, the total number of Hib cases remained very low: only 276 Hib cases were reported among 24 countries in 2021 (Figure 5A). Overall, Hib cases increased among children 0-4 years of age in 2021 (n=146) versus 2020 (n=87; Figure 5C). When stratified by country the increase in Hib disease was primarily observed in five countries: France (6), Israel, Paraguay, South Africa and The Netherlands (14); and there were between 10 and 70 Hib cases in each of those five countries in 2021 (Figure 5E). Among cases of *N meningitidis* there was a significant reduction in disease due to all serogroups (but especially capsule groups W, C and Y), with no obvious changes in the patterns of disease by age group (Figure 5B, 5D).

**Figure 5.**
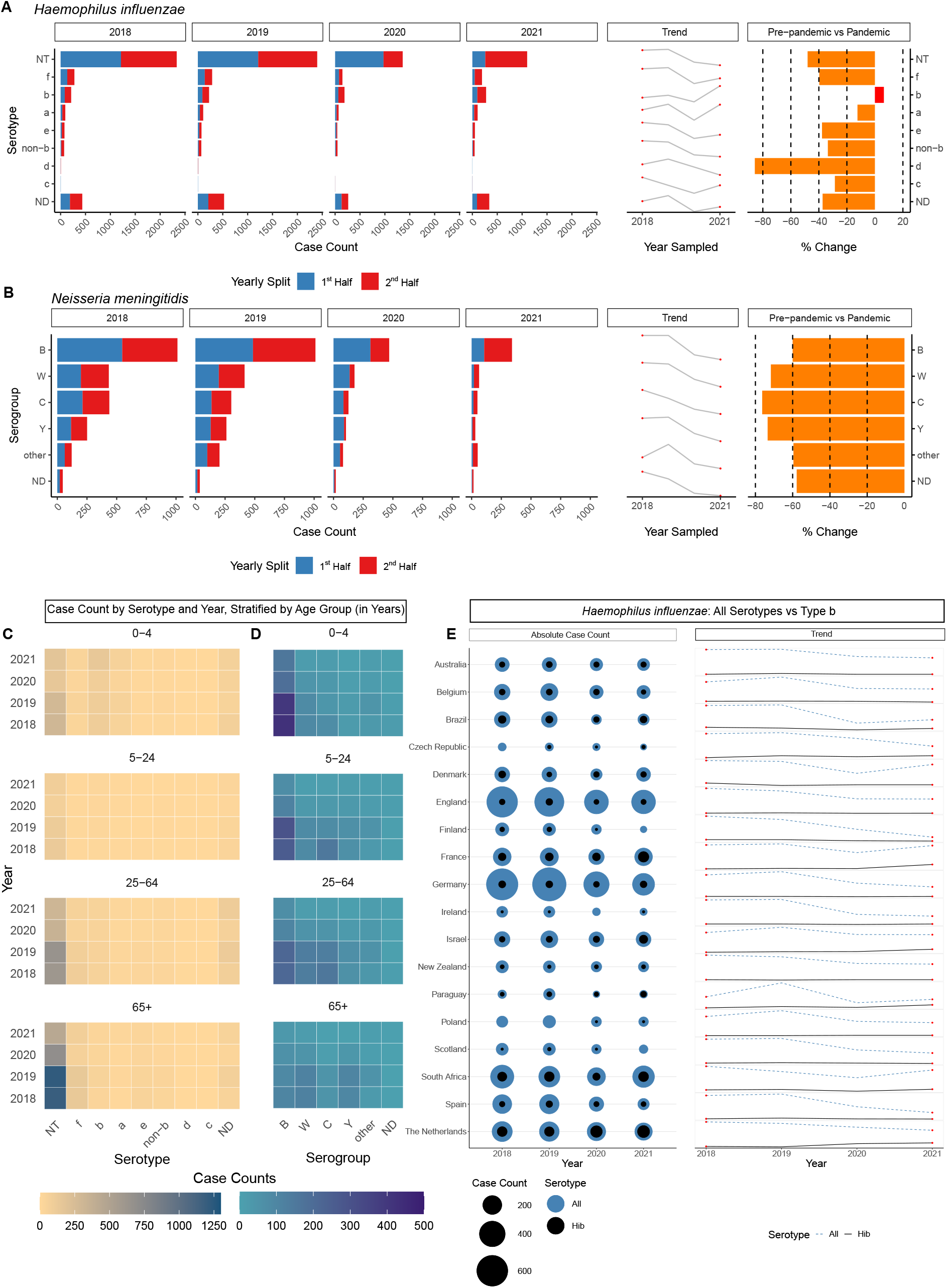
*H influenzae and N meningitidis* invasive disease cases by serotype, capsule group, country and age. Panels A (*H influenzae*) and B (*N meningitidis*): Distribution of serotypes between 2018-2021, listed by case count, increasing or decreasing trend year-by-year, and percentage change of each serotype recovered in 2018-2019 compared to 2020-2021 (average number of cases each year, pre-pandemic vs pandemic). Panels C (*H influenzae*) and D (*N meningitidis*): Heat maps depicting the number of cases of each serotype recovered per year and by age group. Panel E: Number of cases of *H influenzae* by country, displaying only those countries where at least 100 cases in total had been reported across all four study years. Circles represent the total number of cases each year, and lines indicate the increasing or decreasing trend year-by-year, marking ‘All’ serotypes in blue and *H influenzae* serotype b (Hib) in black. Note: serotype ND = not determined.

## Discussion

Our statistical models estimated that more than 36,000 cases of life-threatening invasive bacterial diseases caused by *S pneumoniae, H influenzae* and *N meningitidis* were averted in the countries participating in IRIS during the first two years of the COVID-19 pandemic. The large number of averted cases reduced the morbidity and mortality associated with these infections and would have eased the burden on some healthcare systems during the pandemic. The documented reduction in disease was most plausibly due to the worldwide implementation of COVID-19 containment measures aimed at reducing transmission of SARS-CoV-2, which simultaneously reduced transmission of other microbes that spread via respiratory secretions, including those studied by the IRIS Consortium.

Our findings cannot be explained by underreporting by hospitals and laboratories that were overwhelmed by the pandemic. In most of the countries represented here, it is a legal requirement to report cases of invasive disease caused by the pathogens in this study, and the inclusion of *S agalactiae* as a non-respiratory comparator organism provided reassurance that the surveillance programmes in these laboratories were functioning without major disruptions in the midst of the pandemic.

The current IRIS data demonstrated that whilst cases of disease remained significantly lower in 2020-2021 compared to pre-pandemic levels, rates of disease were increasing in some countries towards the end of 2021 (15,16). It is therefore reasonable to predict that rates of invasive bacterial disease may return to pre-pandemic levels in due course. Furthermore, the 2021 data showed a small overall increase in Hib cases after an initial decrease in 2020. However, there may be other factors that influenced these dynamics even before the pandemic, eg changes to Hib vaccines and/or vaccine schedules, or pre-pandemic increases in the incidence of Hib disease (6,14). Renewed emphasis on the active surveillance of invasive diseases caused by Hib is certainly warranted going forward.

An important concern now, as people move back to normalised social interactions, is which pathogens will cause disease. The usual patterns of microbial transmission were altered during the pandemic, and if the microbiome within the upper respiratory tract was also disrupted (17,18), this could lead to changes in the prevalence of serotypes/groups associated with disease, increased prevalence of circulating nonvaccine types, or emergence of non-traditional disease-associated types. There is also growing concern around decreased population immunity or an ‘immunity debt’ (ie a higher proportion of susceptible individuals within a population because of reduced exposure to commonly circulating microbes) as a result of pandemic restrictions, which could lead to future outbreaks of disease (19,20). Certain populations might be at greater risk of infection, such as children born during the pandemic, teenagers and young adults because of their increased social mixing, and the elderly because of immunosenescence, high rates of underlying comorbidities and frailty. The situation is further compounded by disruptions in routine vaccination schedules around the world since healthcare systems were reorganised to deal with the threat of the pandemic, but at the expense of providing other essential public health services (21-24). Reinstating routine paediatric vaccination programmes is one of the most important post-pandemic challenges the world needs to address as quickly as possible.

In addition to reduced exposure, the typical patterns of respiratory disease were disrupted during the pandemic, and the prevalence of commonly circulating respiratory viruses such as respiratory syncytial virus and influenza viruses were also reduced during the pandemic due to the implementation of COVID-19 containment measures (25,26). Two recent studies reported a correlation between the reduced prevalence of respiratory viruses and reductions in *S pneumoniae* disease during the pandemic (27,28). Further work will be necessary to better understand any causative relationship between respiratory viruses and colonising nasopharyngeal bacteria, mechanisms of co-infections, and mechanistically how one microbe might influence the pathogenicity of another.

Limitations of the time series analyses in this study included difficulties when fitting ARIMA models to individual countries. Each country has its own pattern of disease and public health restrictions, and some countries reported relatively small case numbers. This necessitated temporal and geographical pooling by month and hemisphere respectively, leading to wider prediction intervals and reduced predictive power of the models. Whilst we assessed data from 2018 to 2021, when taking account of a yearly trend, the necessary seasonal adjustment in the model leads to a loss of one year’s worth of data, which impacted our sample size (12).

Despite these limitations, strengths of these analyses included large datasets rapidly contributed by investigators, which spanned four years of time and increased the power of these analyses. Secondly, high quality data at a national level were made available by accredited reference laboratories undergoing routine audit and data validation practices, which minimised information bias. Selection bias is likely to be limited because the bacteria under investigation cause diseases that necessitate urgent hospital care, and legislation in most of these countries mandates the reporting of invasive diseases due to one or more of these bacteria. We also tested a range of time series analyses to ensure robustness of results, and the findings were reproducible.

As our societies emerge from the COVID-19 pandemic, this large prospective study undertaken by the IRIS Consortium allows for timely detection of changes in invasive diseases caused by *S pneumoniae, H influenzae* and *N meningitidis* and provides a means to detect and act on significant changes that will undoubtedly occur. Most importantly, it is essential that any disruptions to bacterial vaccination programmes are resolved since diseases due to these bacteria are devastating but can be prevented by safe and effective vaccines already widely used in many countries worldwide.

## Data Availability

Sharing study data is not possible because of the risk of identifying individual cases of invasive disease in some countries.

## Contributors

ABB, KAJ, MCJM, and MPGvdL conceived the idea for the IRIS Consortium, set up the database infrastructure, and recruited laboratories to join IRIS. ABB, DS and KAJ uploaded, curated and verified data. DS, NDM and ABB analysed and interpreted the data. DS created the figures. DS and ABB wrote the first draft of the paper. All authors reviewed and critically revised the paper for important intellectual content and approved the final version to be published. All authors agreed to be accountable for all aspects of the work. All authors had full access to all the data in the study and the corresponding author had final responsibility for the decision to submit for publication. All authors were involved in the acquisition, processing, or validation of microbiological data, Oxford COVID-19 Government Response Tracker (OxCGRT) data, or Google COVID-19 Community Mobility Reports data.

## Data sharing

Sharing study data is not possible because of the risk of identifying individual cases of invasive disease in some countries. Source code for the statistical analyses will be available via GitHub at the time of publication.

## Acknowledgements

The authors were deeply saddened by the sudden death of Prof Ulrich Vogel, who contributed to the IRIS Consortium but more importantly was a dear friend and colleague to many of the authors.

The authors appreciate the time and effort contributed to IRIS by these individuals: Bianca Crowder and Trang Nguyen (Australia); Samanta Cristine Grassi Almeida, Maria Cristina Brandileone and Ana Paula Lemos (Brazil); Clara Inés Agudelo and Elizabeth Castañeda (Colombia); the clinical microbiology laboratories and technical staff at the Expert Microbiology Unit, THL (Finland); Stelmos Simantirakis and Athanasia Xirogianni (Greece); Wendy Bril-Keijzers (Netherlands); Pilar Ciruela, Juan J Garcia-Garcia, Minako Nagai, Alba Redin, and Liliana Rojas (Spain).

The infrastructure for the IRIS Consortium is funded by a Wellcome Trust Investigator Award to ABB (grant number 206394/Z/17/Z). The IRIS databases are part of PubMLST, which is funded by a Wellcome Trust Biomedical Resource Grant awarded to MJCM, ABB, and KAJ (grant number 218205/Z/19/Z). DS is funded by an Oxford Clarendon Scholarship. The high-performance computing requirements of the data analyses were supported by the Wellcome Trust Core Award (grant number 203141/Z/16/Z) and the NIHR Oxford Biomedical Research Centre. The views expressed are those of the authors and not necessarily those of the NHS, the NIHR or the Department of Health. Authors were not precluded from accessing data in the study, and they accept responsibility to submit for publication.

## Notes

### Competing Interest Statement

The authors have declared no competing interest.

### Author Declarations

A waiver of ethical approval was received from the Chair of the Oxford Ethical Committee (Dr. Kevin Marsh).

