## Supplementary figures and images for "Sustained reductions in life-threatening invasive bacterial diseases during the first two years of the COVID-19 pandemic: analyses of prospective surveillance data from 30 countries participating in the IRIS Consortium"

### Supplementary Figure 1

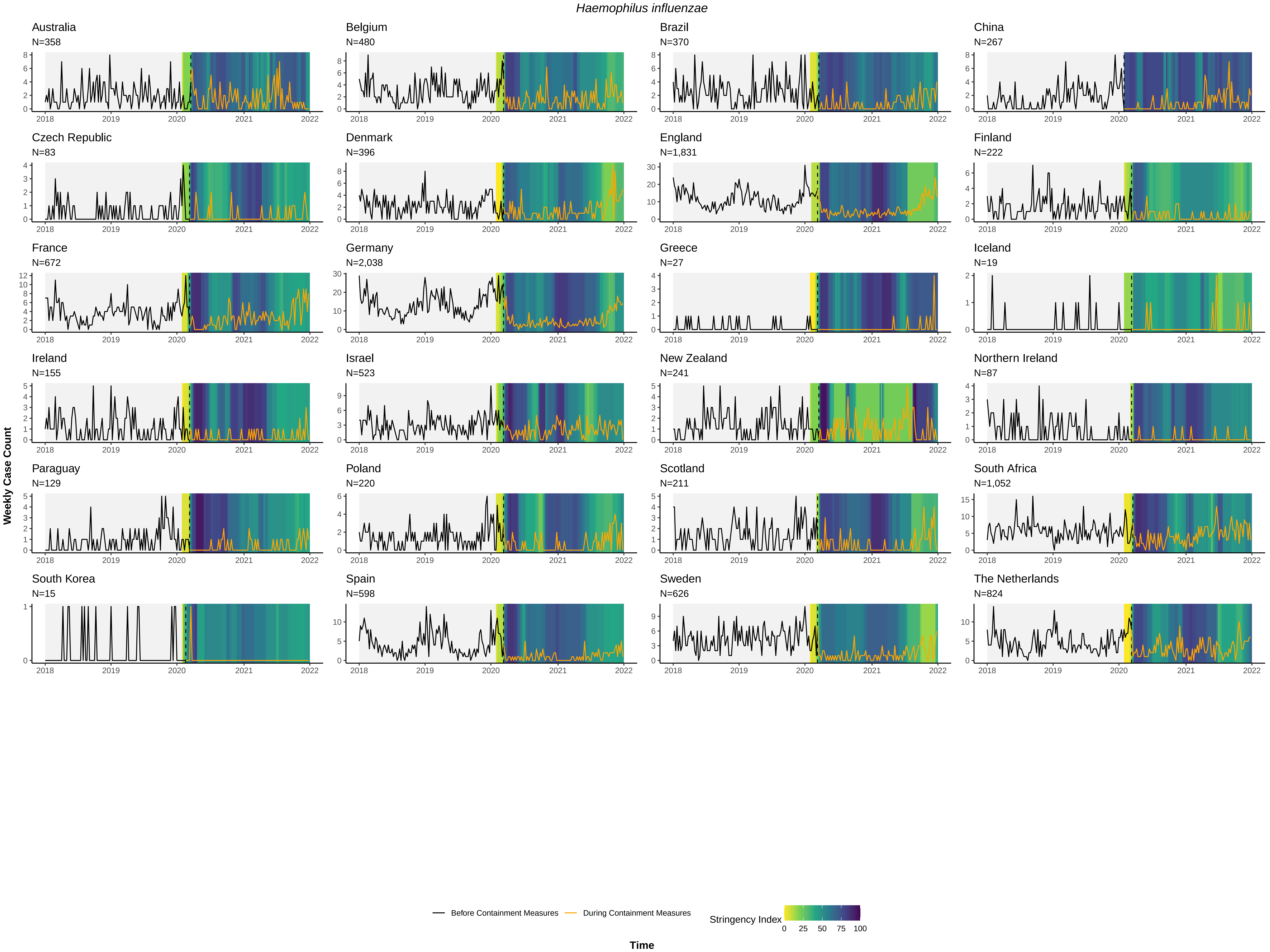

### Supplementary Figure 2

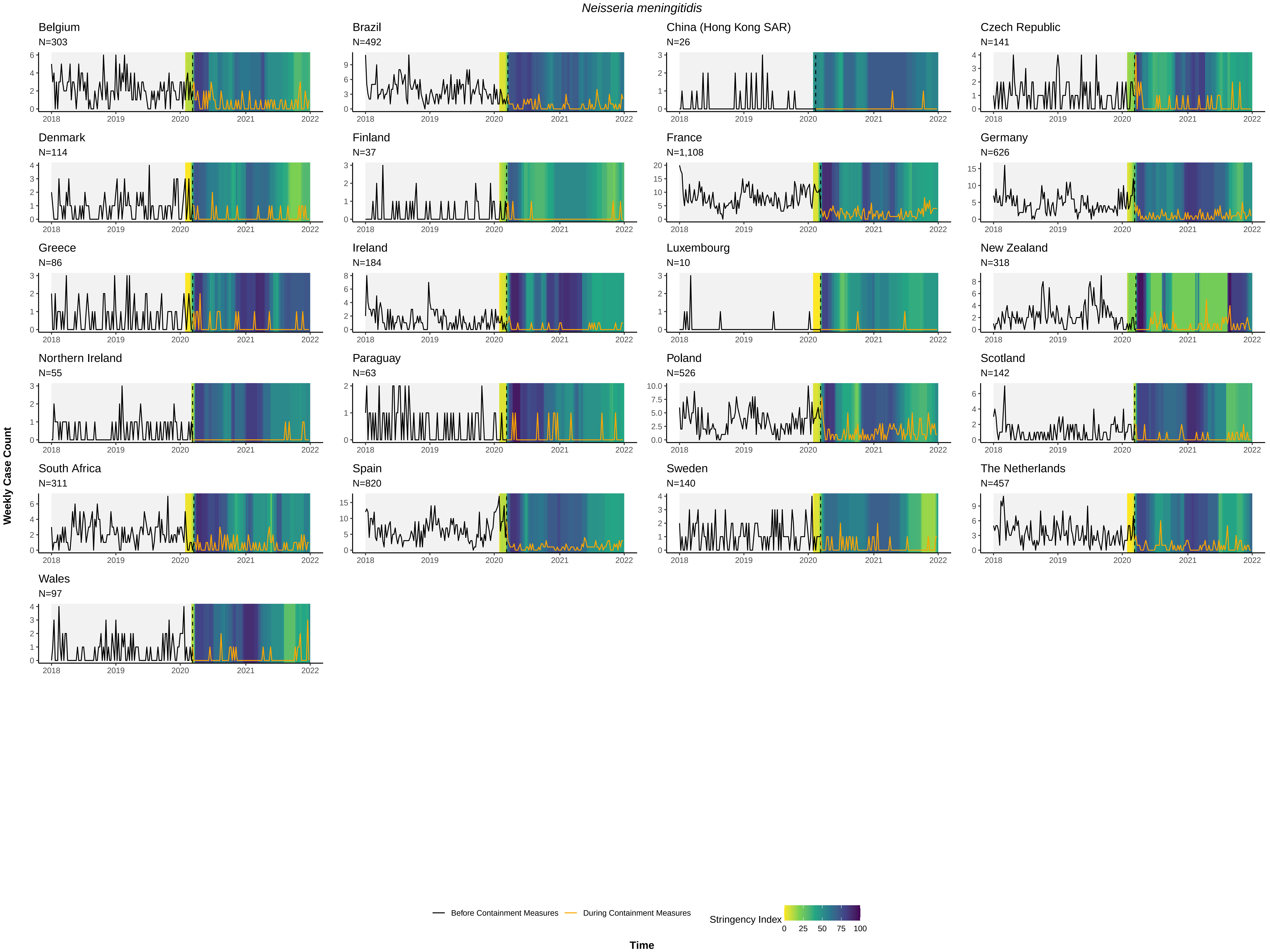

### Supplementary Figure 3

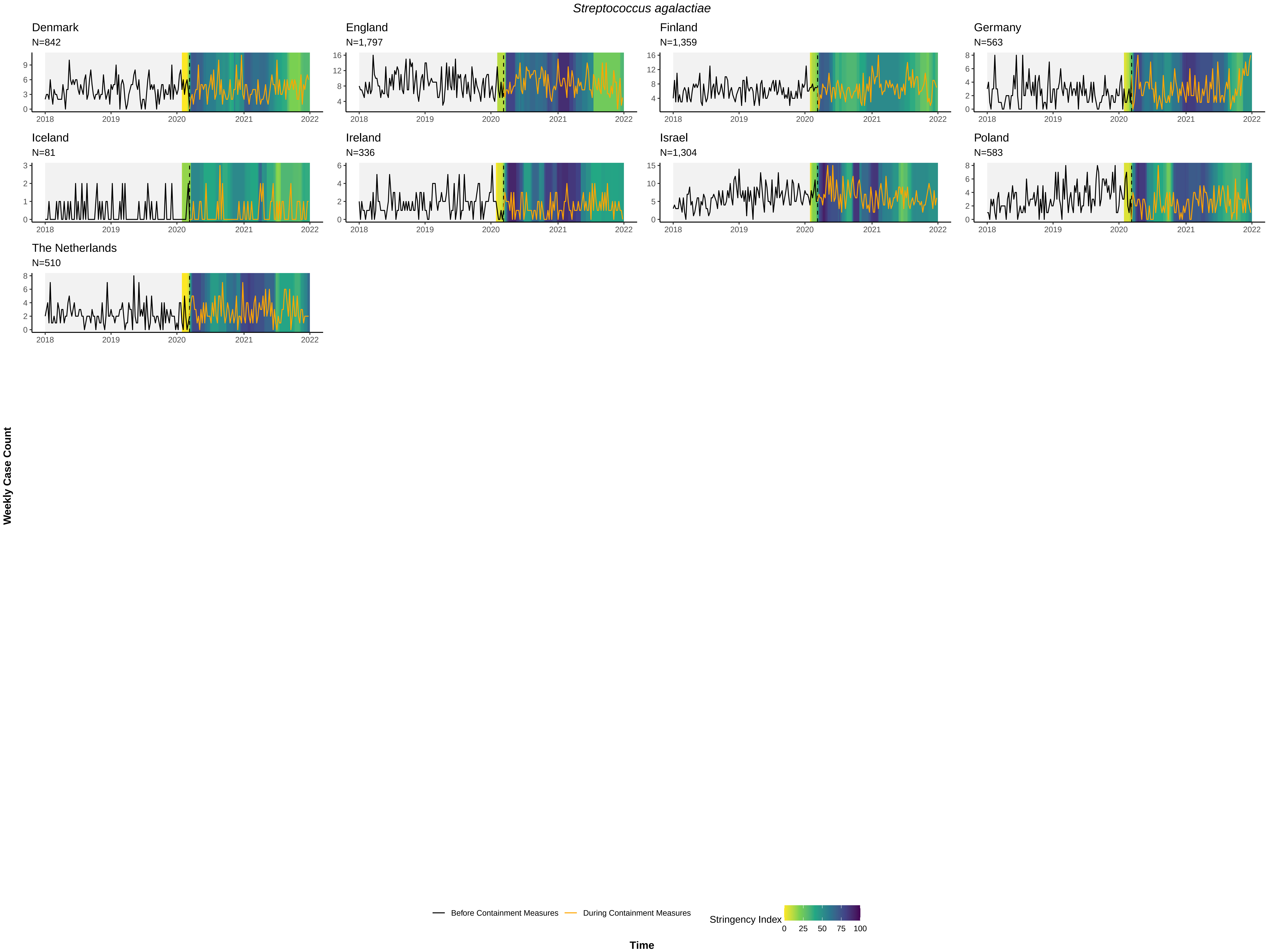

### Supplementary Figure 4

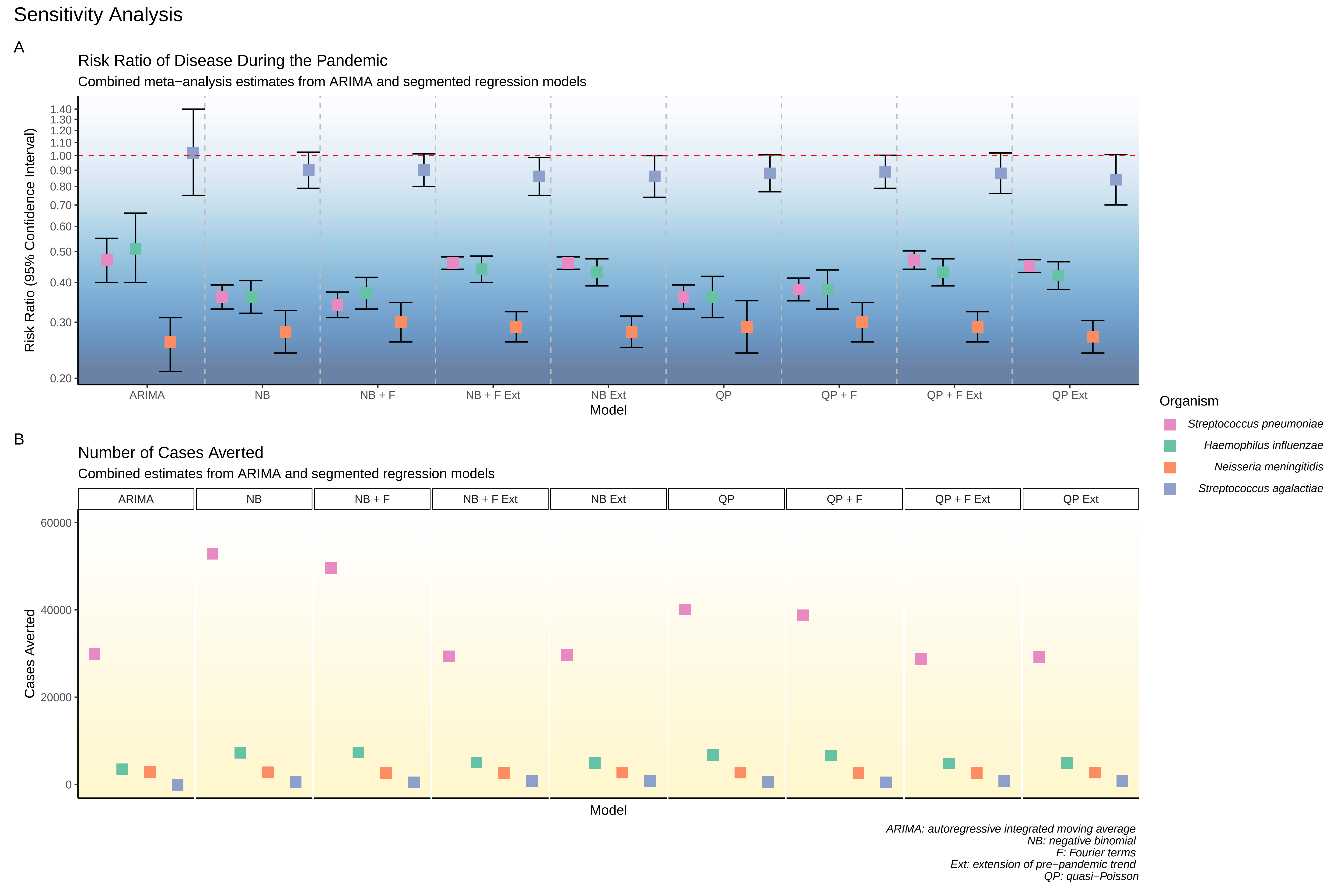
